# Exploring provider perspectives on patients with atrial fibrillation who are not prescribed anticoagulation therapy

**DOI:** 10.1101/2021.03.09.21253217

**Authors:** Catherine Yao, Aubrey E. Jones, Stacey Slager, Angela Fagerlin, Daniel M. Witt

## Abstract

**Background:** Evidence-based guidelines strongly recommend oral anticoagulants to prevent stroke in patients with non-valvular atrial fibrillation (AF). However, many patients are not prescribed guideline recommended anticoagulant therapy.

**Objectives:** To explore themes underlying anticoagulant prescribing or discontinuation in patients with AF and compare and contrast prescribing preferences between cardiologists and general practitioners.

**Methods:** Providers at the University of Utah Health system were recruited to participate in semi-structured interviews. An interview guide directed the 15-minute interviews with focus on anticoagulant prescribing practices for patients with AF. Interviews were transcribed verbatim. Two reviewers independently read transcripts and labeled passages of text corresponding with key concepts and themes.

**Results:** Of the eleven practitioners interviewed, seven practiced in cardiology, two in internal medicine and two in family medicine. The most prominent reasons cited for not prescribing anticoagulation for stroke prevention in AF patients were concerns about intracranial bleeding, followed by gastrointestinal bleeding. Other common reasons were increased age, thrombocytopenia, chemotherapy, previous or concerns of noncompliance, and comorbidities. Providers believed patient refusal of anticoagulants was related to fear of bleeding, medication burden, or warfarin’s negative reputation. All prescribers reported similar prescribing strategies, including using risk stratification, shared decision making, and utilizing specialized anticoagulation clinics and pharmacists as resources.

**Conclusion:** Fear of bleeding was a common theme underlying anticoagulant underutilization in patients with AF. Identifying major reasons directly from providers can be utilized to develop patient education addressing common fears and misconceptions, promote shared decision making, and provide provider education and resources to achieve appropriate anticoagulant prescribing.

## Introduction

One in four Americans are diagnosed with atrial fibrillation (AF) sometime in their lifetime, making it the most common arrhythmia type.^1^ AF is known to increase the risk of stroke by three to five-fold due to blood stasis and hypercoagulability caused by irregular contraction in the left atrium.^2^ Extensive data from randomized controlled clinical trials support the efficacy of anticoagulants in reducing stroke risk in patients with non-valvular AF.^3,4^ However, many eligible patients are not prescribed anticoagulant therapy despite current evidence-based guideline recommendations, and even those who are prescribed often do not take it.^3,5,6^

A commonly used stroke risk stratification scheme for non-valvular AF is the CHA_2_DS_2_-VASc score.^7^ This score assesses the risk of stroke based on the following risk factors (one point per risk, exceptions noted): congestive heart failure; hypertension; age 65-74 years; age over 75 years (two points); diabetes mellitus; prior stroke or transient ischemic attack (two points); vascular disease; and female sex.^8^ Anticoagulant agents, including direct-acting oral anticoagulants (DOACs) or warfarin are recommended for all patients with CHA_2_DS_2_-VASc scores of 1 or more for men and 2 or more for women.^7^ The CHA_2_DS_2_-VASc tool has been historically underutilized in clinical practice leading to missed opportunities in identifying patients who would benefit from anticoagulant therapy.^1,6,9^

Even in patients with CHA_2_DS_2_-VASc scores indicating need for stroke risk reduction, some providers fail to prescribe anticoagulants based on perceived risks such as bleeding, fall risk, or relative contraindications such as renal or hepatic dysfunction or the presence of interacting medications.^3^ Various bleeding risk-stratification tools have been proposed to predict bleeding risk while on anticoagulants but these have limited utility in determining the risk/benefit of anticoagulation therapy and guidelines recommend against using these tools.^7^ The clinical reasoning behind the decision to forgo anticoagulant therapy often goes undocumented in the medical chart.^3^ Inadequate stroke prevention remains prevalent as one-third of patients hospitalized following an AF-related stroke in one study were not receiving anticoagulation therapy despite the presence of stroke risk factors.^3^ Thus, there is a critical gap in understanding the underlying reasons why providers do not prescribe anticoagulants in patients with AF.

The objective of this qualitative study was to explore themes underlying why anticoagulants are under-prescribed from the provider’s perspective and describe general characteristics of patients with AF in whom providers do not feel comfortable prescribing anticoagulant therapy.

## Methods

This qualitative study included interviews with a sample of providers practicing within the University of Utah Health (UHealth) system. The UHealth system encompasses four hospitals and 12 health clinics in Utah. To obtain a range of clinical perspectives, the inclusion criteria consisted of healthcare providers who regularly see patients with AF and have prescribing authority. Participants were chosen from different disciplines, including cardiology, internal medicine, and family practice, and included medical doctors, physician assistants, and nurse practitioners. Providers were invited to participate in interviews and completed the informed consent process through a standardized email.

This study was approved by the University of Utah Institutional Review Board (IRB). Interviews occurred between July and November 2019. A semi-structured interview template (see Appendix) was used to guide the interview and provide consistency. An expert in qualitative research (SS) conducted interviews either in person or by telephone. Interviews lasted, on average 15 minutes.

Provider demographic and practice-related variables including years in practice, medical specialty, and experience with managing AF, strokes, and bleeding in daily practice, were collected. Audio recordings from each interview were transcribed verbatim using Transcribe, a transcription software (Wreally LLC, Los Angeles, CA). Common themes were determined by utilizing an inductive approach where transcribed text was coded and linked to a theme. Two reviewers read the transcripts line-by-line and labeled passages of text to correspond with key concepts. Coding took place independently, but regular meetings occurred to ensure agreement and allow for reconciliation of discrepancies. A major theme for why anticoagulants are not prescribed was established when key concepts were repeated throughout interviews. The number of interviews was determined by thematic saturation, which occurred when major themes became established and no new themes emerged with further interviews.^10,11^

## Results

A total of 11 interviews were conducted before thematic saturation was achieved. Baseline provider demographic and practiced-related information is summarized in Table 1. Most participants were medical doctors (72.7%) with additional input from a physician assistant, advanced practice registered nurse, and nurse practitioner. The different specialties represented were cardiology (63.6%), family medicine (18.2%), and internal medicine (18.2%). Common themes identified from the interviews are listed in Table 2 with supporting quotes (Q) and additional quotes are listed in the appendix.

**Table 1.** Demographic and Clinical Practice Characteristics of Providers

| Characteristics | Providers (N=11) |
| --- | --- |
| **Sex, n (%)** |  |
| Female | 6 (54.5) |
| **Training, n (%)** |  |
| Medical doctor | 8 (72.7) |
| Physician assistant | 1 (9.1) |
| Advanced practice registered nurse | 1 (9.1) |
| Nurse practitioner | 1 (9.1) |
| **Specialty, n (%)** |  |
| Cardiology | 7 (63.6) |
| Family medicine | 2 (18.2) |
| Internal medicine | 2 (18.2) |
| **Practice site, n (%)** |  |
| Inpatient | 1 (9.0) |
| Outpatient | 5 (45.5) |
| Both inpatient and outpatient | 5 (45.5) |
| **Length of practice (years)** |  |
| Mean (standard deviation) | 5.96 (±6.18) |
| **Number of atrial fibrillation patients seen per week** |  |
| Mean (standard deviation) | 8.72 (±5.10) |

**Table 2.**
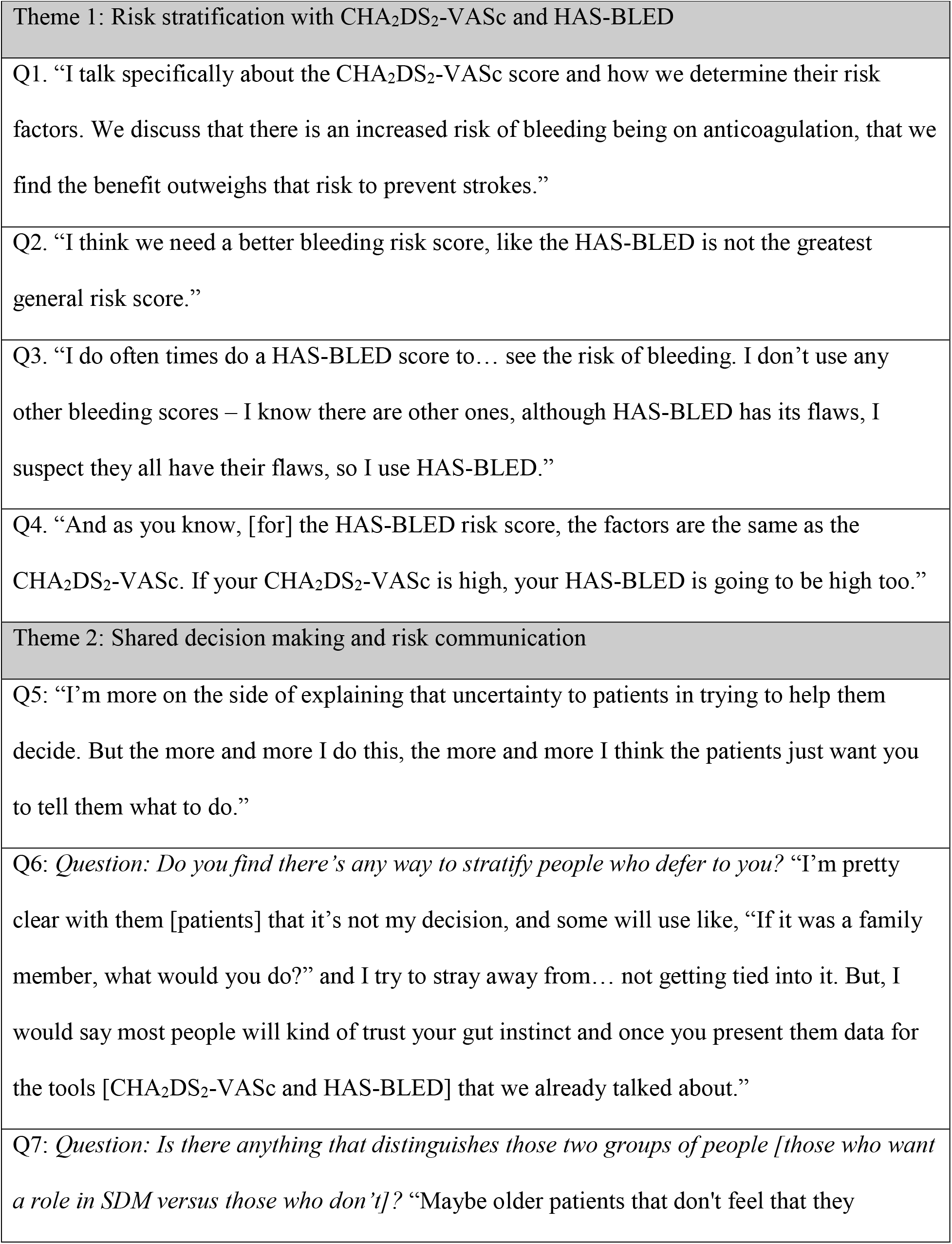
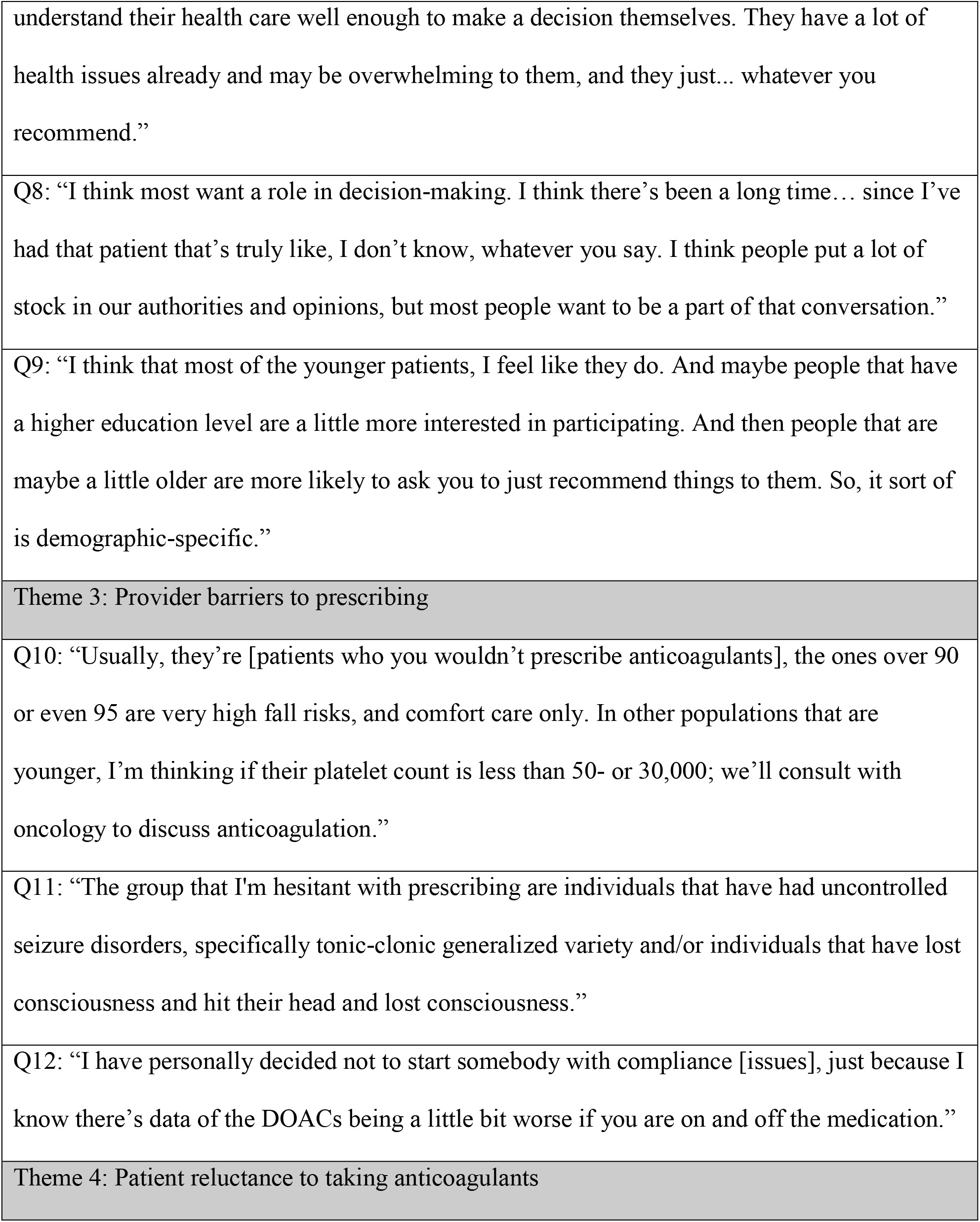
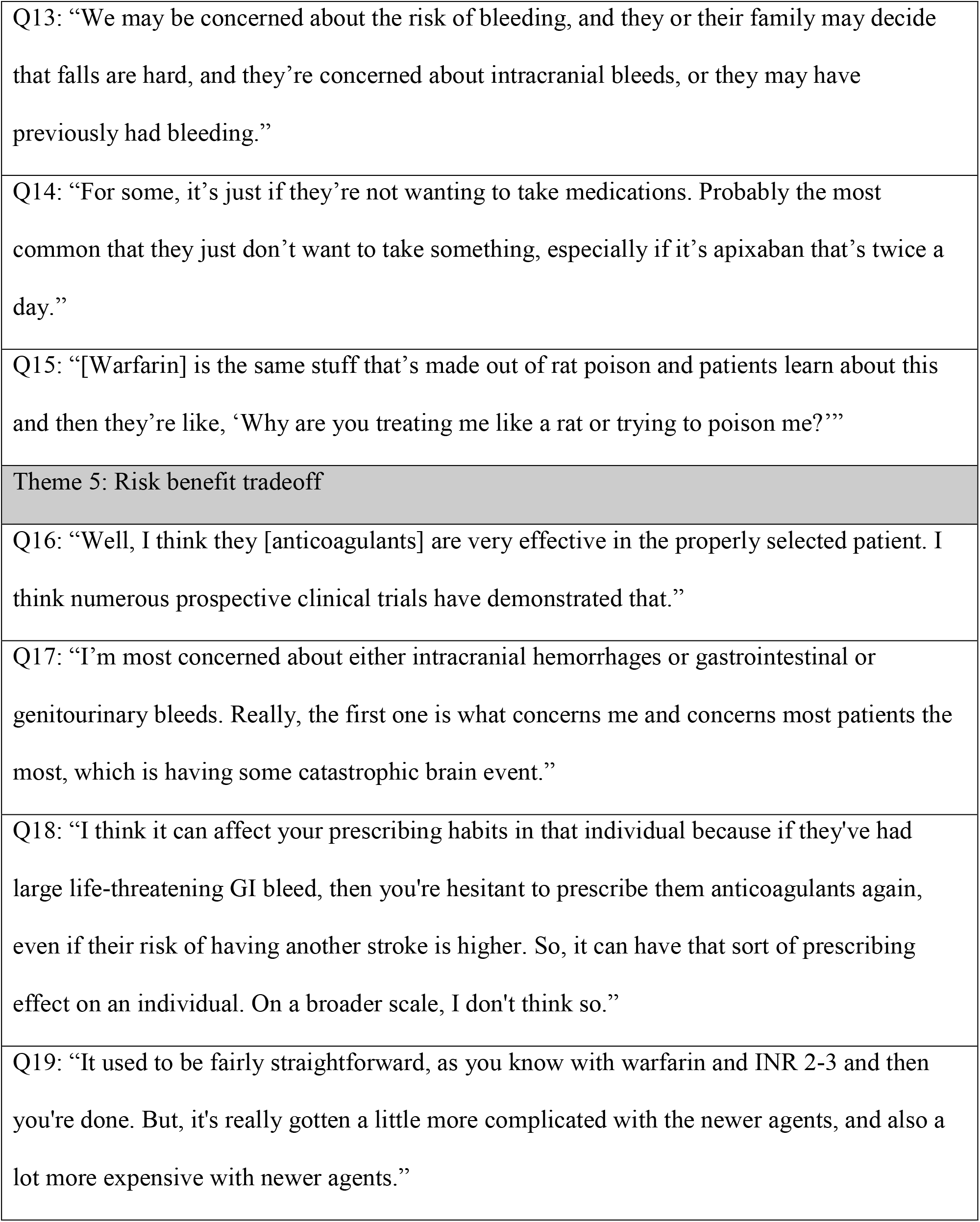
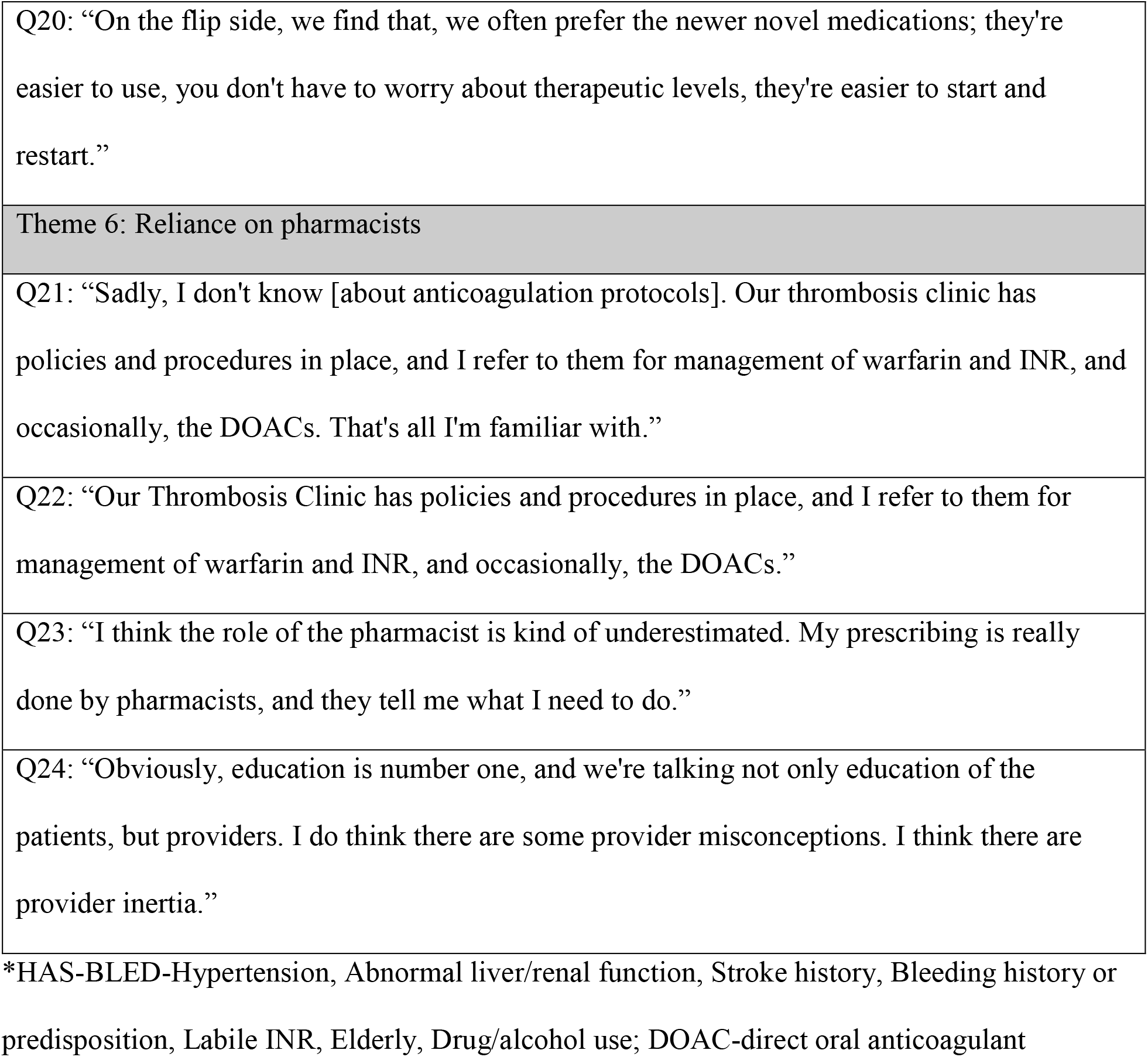
Selected Themes and Supporting Quotes

### Risk Stratification

A common theme was providers utilizing both the CHA_2_DS_2_-VASc and bleeding risk scores to stratify the stroke and bleeding risk for a given patient, respectively (Q1). Ten of eleven providers reported using CHA_2_DS_2_-VASc for every AF patient as part of their routine. Almost half of the providers also mentioned using a bleeding risk score, although they also acknowledged flaws with these tools (Q2-4).

### Shared Decision Making

All providers reported discussing the risk/benefit of stroke prevention vs. bleeding with their patients before prescribing anticoagulants. Many providers had a similarly structured conversation for counseling patients with AF, including the predicted risk of stroke and how it can be mitigated with anticoagulants (Theme 2). Most providers agreed that starting anticoagulants is a shared decision and try to do some degree of shared decision making with their patients before prescribing. However, all providers reported perceiving varying levels of decision making patient engagement (Q5). Providers believe many patients want a role in the decision making process, although some simply follow any provider recommendations (Q6-8). Specifically, older age and lower education or health literacy levels were perceived as factors that led providers to forgo shared decision making attempts (Q7, Q9).

### Reasons for Not Starting/Discontinuing Anticoagulants

Overall, common barriers from a provider perspective for not prescribing or discontinuing anticoagulants included increased patient age, thrombocytopenia or chemotherapy, noncompliance, and specific comorbidities, such as diabetic neuropathy, seizure disorders, or liver or kidney disease (Q10-12). When asked why some patients refuse to take anticoagulants, or were reluctant to start, providers suggested the most common reasons were patients’ fear of bleeding, medication burden, and the reputation of warfarin, specifically its association with rat poison (Q13-15).

### Risk/Benefit Tradeoff

Ten providers cited randomized clinical trials supporting the benefit of anticoagulants in reducing the risk of AF-related stroke (Q16). In contrast, intracranial bleeds were mentioned by ten providers as the most concerning adverse event, followed by GI bleeds, which was cited by six providers (Q17). The majority stated that GI bleeds are easier to manage, and patients can usually restart anticoagulants. As a result, most providers said that they may be more cautious for an individual with a history of bleeds, but they do not believe it affects their overall prescribing habits (Q18). When describing barriers for prescribing different agents, they mentioned the inconvenience of warfarin, including the labor-intensive monitoring and dose adjustments, as well as the cost and insurance barriers with DOACs (Q19-20).

### Resource Utilization and Areas for Improvement

Most providers were unaware of any institutional protocols or procedures providing guidance for anticoagulant prescribing in patients with AF (Q21). Five clinicians referred their patients to the Thrombosis Clinic, which is a pharmacist-run referral service for management of anticoagulation therapy, as part of their usual process for managing anticoagulants (Q21-22). Additionally, four providers cited pharmacists as a key resource when choosing an anticoagulant (Q23). There were also multiple suggestions for increased provider education, specifically relating to DOACs and keeping up with new evidence and literature in this field (Q24).

## Discussion

This qualitative study explores themes regarding factors that providers report influenced their anticoagulant prescribing decisions for stroke prevention in certain AF patients. In previous studies, bleeding risk was the most common reason providers did not prescribe anticoagulants, followed by history of adverse events and older age.^3,6^ Other important patient factors that were considered were inconvenience of therapeutic monitoring, cognitive dysfunction, impaired renal or hepatic function and patient refusal.^6,9^ This study reports similar results, but provides more insight on the provider’s prescribing process, including shared decision making with the patient and risk vs. benefits analysis.

Based on the common themes emerging from the provider interviews, there are certain areas that can be targeted to increase guideline-recommended anticoagulant prescribing. Specifically, areas of focus are (a) addressing patient-specific fears and misconceptions; (b) encouraging shared decision making; and (c) providing education and promoting interprofessional collaboration as resources for providers.

### Address Patient-Specific Fears and Misconceptions

A common theme for not prescribing anticoagulants was patient refusal or perceived refusal, namely due to a fear of bleeding, increased medication burden, and a negative reputation associated with warfarin. This is an opportunity to increase patient education and utilize motivational interviewing techniques, such as addressing the common myth that prescription warfarin is rat poison. Patients may also require more counseling focused on the perceived risk of bleeding compared to the actual risk. One provider discussed how bleeding is usually rare and treatable, while stroke is potentially devastating, so she tends to err on the side of using anticoagulants.

### Shared Decision Making

While all providers discussed the risks and benefits of taking anticoagulation for stroke reduction, this is only one aspect of shared decision making.^12^ Most providers affirmed that they engaged in shared decision making with most of their patients, however, it is unclear whether shared decision making actually occurs as studies have shown that patient involvement in decision making is not consistently taking place.^13,14^ Our findings are in line with previous studies showing that providers perceive that certain types of patients are less likely to want to engage in shared decision making (i.e. age and education level), however other studies have shown that these patient factors are not accurate predictors of engagement in shared decision making.^15-18^ Future studies with validated outcome measures and recordings of provider-patient interactions could help clarify how prevalent actual shared decision making is when starting anticoagulants.

### Provide Education and Promote Interprofessional Collaboration as Resources for Providers

As ten out of 11 providers reported using the CHA_2_DS_2_-VASc score to stratify stroke risk with virtually every AF patient, it is apparent that the score is utilized appropriately as recommended by evidence-based guidelines. The only provider who rarely used CHA_2_DS_2_-VASc worked in family medicine and usually managed patients already on anticoagulants. Four cardiologists mentioned the HAS-BLED to stratify bleed risk, while the atherosclerotic cardiovascular disease (ASCVD) risk calculator was used by two family medicine providers. This distinction may be used as context for future provider education – cardiologists may have a primary goal of assessing stroke vs. bleeding, while PCPs may have a target of decreasing atherosclerotic cardiovascular events in general.

Although providers are most concerned with intracranial hemorrhages, the majority shared a similar perspective that critical bleeds are very infrequent. A few providers noted that the fragmented healthcare system may contribute to a perceived lower risk of bleeding or stroke. Since treating an emergent bleed or stroke is out of the scope of practice for general cardiologists or PCPs, they generally do not see these traumatic events. This perspective suggests that an interprofessional approach to patient care and closed healthcare delivery systems may improve prescribing practices and coordination of care.

As a majority of providers were unaware of institutional prescribing guidelines, all providers would benefit from being updated on local recommendations and resources by clinical anticoagulation services such as the Thrombosis Clinic. Multiple providers mentioned utilizing pharmacists to stay up to date on evidence-based guidelines, choose a specific anticoagulant, manage INR results, and help with cost-conscious prescribing. Based on these answers, pharmacists have an opportunity to promote their clinical services for providers, as well as provide more system-wide education on current literature and cost. Promoting overall interprofessional collaboration can result in comprehensive, evidence-based, and patient-centered anticoagulation management.

By interviewing providers directly, this qualitative study evaluated providers’ perspectives on clinical decision-making and which patient-specific factors are considered. Gaps in the prescribing process where targeted interventions can be made were highlighted. The results can be used to generate future quantitative studies on the subject. A limitation of the study is the small number of interviews conducted within one healthcare system; the answers and themes may change across other providers and healthcare systems, specifically if the institution does not have a specialty anticoagulation clinic. Additionally, it cannot be concluded that providers were accurate reporters of their behaviors. As is true for all qualitative studies, we generated potential hypotheses related to anticoagulant prescribing for patients with AF but these hypotheses require testing in quantitative studies.

## Conclusion

The goal of this qualitative study was to identify themes relating to prescribing anticoagulants for patients with AF. Identified themes can be utilized to implement targeted interventions and provider education aimed at increasing guideline-directed prescribing. There are common patient fears that leads to anticoagulant refusal, suggesting improved patient education, counseling, or support groups that address specific misconceptions may reduce the likelihood that patients refuse anticoagulants. Most providers interviewed indicated that they utilized discussions of the risk vs. benefit of starting an anticoagulant and that the decision to initiate anticoagulation therapy should involve the patient. Conversely, some providers have the perception that patients may not want to engage in shared decision making or that they are less likely to engage in shared decision making with some patients. Future studies with validated shared decision making tools and outcome assessment are needed to quantify the benefit of shared decision making when prescribing anticoagulants. Lastly, it may be beneficial to focus provider-education on institutional policies, referral services for anticoagulation management, and interprofessional collaboration with pharmacists. Focusing future interventions on the themes identified in our study will help increase our chances of improving outcomes for patients with AF.

## Supporting information

Interview Questions and Supplemental Table 1

## Data Availability

Data is available within the article. Further data may be requested from the authors.

