## Supplementary material for "Exploring provider perspectives on patients with atrial fibrillation who are not prescribed anticoagulation therapy": Interview Questions and Supplemental Table 1

**Appendix**

Standardized Interview Template

Introduction

1. What is your practice specialty? Interventional or generalist? Inpatient or outpatient?
    a. How long have you been practicing?
    b. Do you treat patients with active or a history of bleeding or stroke?
    c. Do you have any research experience in this field?
2. How many patients with AF do you usually see on a weekly basis?
    a. Do you perform procedures for AF (e.g. Watchman, ablation)?
3. How confident do you feel about prescribing and managing anticoagulants?
4. Do you have any personal/family experiences with AF or stroke?

Clinical approach

5. How often do you use $CHA_2DS_2VASc$ to stratify stroke risk in patients with non-valvular AF? If you don't generally use $CHA_2DS_2VASc$, what is your approach to determining stroke risk?
6. What protocols/policies are in place at your practice setting on prescribing anticoagulants in AF patients? How often do you deviate from the protocol/policy? What are some common reasons for not following the protocol/policy?
7. Tell me about the typical conversations you have with patients with AF before starting anticoagulants?
    a. What proportion of patients refuses to take anticoagulants? What are common reasons why patients refuse to take anticoagulants?

b. What role do patients want in this decision-making? Or do you just make the clinical recommendation?

Reasons for not prescribing anticoagulants

8. What are your thoughts on the effectiveness of anticoagulants (DOACs, warfarin) for preventing stroke in patients with AF?
9. What adverse events are you most concerned about? How frequent are they? How does that affect your prescribing habits?
10. What type of patient is most commonly *not* started in anticoagulation despite potential eligibility?
    a. Why don't you anticoagulate them?
    b. What are some absolute contraindications for anticoagulants?
11. What are some common reasons why you discontinue anticoagulants?
12. Is there anything else you would like me to know about prescribing anticoagulants to prevent stroke in patients with AF?

Additional Supporting Quotes

| Theme 1: Risk stratification with $CHA_2DS_2$-VASc and HAS-BLED |
| --- |
| "Virtually, always [use $CHA_2DS_2$-VASc] . There's some cases where anticoagulation would be relatively contraindicated and I wouldn't bother, perhaps. Somebody with a history of recurrent catastrophic bleeding, for example, on dialysis who's never had a stroke – somebody like that, I would probably not even bother calculating. But otherwise, yes, it's pretty much a routine." |
| "As you know, the HAS-BLED risk score – the factors are the same as the CHA2DS2-VASc. If your CHA2DS2-VASc is high, your HAS-BLED is going to be high, too. So, a lot of this, at the end of the day, is art applied to science and science applied to art." |

| Theme 5: Risk benefit tradeoff |
| --- |
| "Traumatic head bleed and GI bleeding. GI bleeding… I would say once a month, we'll have someone that needs to stop their anticoagulation for most commonly a slow GI bleed so it's not emergent. They don't have to go to the ER; it does not require blood transfusions, but they need to have some work-up and then they're able to restart it. Occasionally, I'd say less frequently, and I can't recall of any specific recent cases of an emergent GI bleed or head bleed so those are rare." |
| "I think the only one that is always the huge concern, although obviously infrequent, is the intracranial hemorrhage. Because GI bleeding is more frequent, that probably gets more discussion. But the one that I worry most about obviously is intracranial hemorrhage." |

| Theme 6: Reliance on pharmacists |
|---|
| "I really appreciate the role of pharmacists with this because most of the newer agents weren't out when I was a student, and so in trying to figure out how to incorporate those into current practice, having a pharmacy team to help has been really helpful. They're my favorite!" |
